# Metagenomic detection of Mimiviridae reads in upper respiratory tract samples of SARS-CoV-2 patients

**DOI:** 10.1101/2024.09.17.24313786

**Authors:** Siddharth Singh Tomar, Krishna Khairnar

**Author notes:** Corresponding author: Correspondence to Krishna Khairnar.

## Abstract

The upper respiratory tract (URT) virome is crucial in respiratory health and response to pathogens. While common respiratory viruses are well-studied, the presence and potential impact of giant DNA viruses, such as Mimiviridae, in the human URT remain underexplored. This study employed a whole genome metagenomics approach to profile the URT virome of 48 SARS-CoV-2-positive patients from central India. Mimiviridae reads were detected in two elderly male patients with severe acute respiratory infection (SARI) or influenza-like illness (ILI), contributing to 24% and 44% of the total virome in their samples. The dominant species were *Acanthamoeba polyphaga mimivirus* and *Moumouvirus*. Although Mimiviridae are not traditionally associated with human respiratory infections, their presence in SARS-CoV-2 patients raises questions about their potential role in co-infections and disease severity, particularly in individuals with ongoing respiratory infections. These findings underscore the need to investigate further giant viruses’ clinical significance, transmission, and pathogenicity in humans. Future research should focus on their epidemiology and the development of improved diagnostic tools to assess their contribution to human health.

## 1. Introduction

The upper respiratory tract (URT) virome has an essential role in the respiratory health of humans. Depending on the context, the virome interacts with the host’s immune system and could either enhance or dampen the host’s immune responses to pathogenic viruses^1^. Certain viruses that are otherwise part of normal virome during a respiratory infection can exacerbate conditions like asthma and chronic obstructive pulmonary disease (COPD)^2,3,4^.

Therefore, it is crucial to understand the dynamics of the URT virome and its interactions with the host immune system and other constituents of the URT microbiome, such as bacteria and fungi. Common respiratory viruses and their role in the URT microbiome are adequately explored through Metagenomics, qRTPCR and viral culture-based studies^5,6,7,8^. However, giant DNA viruses like Mimivirus have also been reported in human respiratory samples suspected of respiratory illness ^9,10,11,12^. Studies about giant viruses are generally limited due to the initial limitations of molecular detection and culturing^13^.

Therefore, the potential role of these giant viruses in the URT microbiome has yet to be thoroughly investigated. Several researchers have used metagenomic approaches to overcome these limitations^14,15,16,^ as the metagenomic approach provides a holistic insight into compositional and functional aspects of URT virome. Since its unexpected discovery in 1992 in Bradford, England, among the samples from a cooling tower during a pneumonia outbreak investigation^17^, Mimivirus has traditionally been associated with respiratory illness. However, the evidence for this association is relatively sparse and conflicting; there are *in Vivo* studies in mice models suggesting the role of Mimivirus in inducing pneumonia^18^ in addition to this, there are studies where seroconversion against Mimivirus is reported in humans ^19,20,^ contrary to this there are studies ruling out the causative association of Mimivirus with pneumonia ^21.^ Pneumonia is also a common manifestation of respiratory illness caused by SARS-CoV-2 infection; therefore, it is imperative to investigate the presence of Mimivirus in COVID-19 patients and their potential role in disease outcomes. Testing laboratories worldwide still have large repositories of URT swab samples that were collected for SARS-CoV-2 molecular testing and genome surveillance during the COVID-19 pandemic. These samples may serve as a starting point for large-scale studies investigating the composition of URT virome using a metagenomic approach.

Therefore, we have decided to retrospectively investigate the URT samples using untargeted metagenomics. These samples were received initially at our lab for SARS-CoV-2 genome surveillance from districts of the Vidarbha region of central India. In this study, we have used the untargeted metagenomics-based approach for profiling the URT virome of SARS-CoV-2 patients from central India. The metagenomic data revealed some interesting facets of the URT microbiome in SARS-CoV-2 patients, including detecting Mimivirus DNA reads from URT samples of SARS-CoV-2 patients.

## 2. Materials and methods

The materials and methods were as per Tomar and Khairnar 2024 ^22^. Briefly, the modifications to the methodology used for this work are as follows: the threshold of nucleotide (NT) reads per million (rPM) ≥ 5, and alignment length ≥ 35 was used, Krona charts were generated from Centrifuge^23^ classification data using the Commander bioinformatics platform^24^, and CZid*s taxonomic tree view feature was used for taxonomic tree visualisation.

The sequencing data were analysed using the Chan Zuckerberg ID (CZid) software. ERCC sequences were removed with Bowtie2, and further filtering was done with fastp to eliminate adapters, low-quality reads, short reads, low-complexity regions, and reads with undetermined bases. Human DNA reads were removed using Bowtie2 and HISAT2, and duplicate sequences were filtered out using CZid-dedup. Using GSNAP, RAPSearch, Minimap2, and the Diamond tool, non-human reads were aligned to the NCBI nucleotide (NT) and protein (NR) databases. Short reads were assembled into contigs using SPADES, and contig-read associations were restored with Bowtie2. BLAST analysis was performed on contigs against the NT and NR databases.

The nasopharyngeal and oropharyngeal (NP-OP) swab samples were collected in Viral Transport Medium (VTM) by trained staff at healthcare centres and SARS-CoV-2 testing labs in the Vidarbha region of Maharashtra, India. Aliquots of samples were stored at -80°C, and 48 samples were selected for whole-genome metagenomics (WGMG) sequencing. The participants had a median age of 36 years (IQR: 16-65), with half of the samples from females.

The DNA extraction was performed using the QIAamp DNA Microbiome Kit, and quality control was carried out with Qubit and Nanodrop. Libraries were prepared with the QIAseq FX DNA Library Kit, and sequencing was conducted on the NextSeq 550 platform.

## 3. Results

Mimiviridae reads were detected in 2 (S3 & S21) out of 48 upper respiratory tract samples obtained from SARS-CoV-2-positive patients. These two samples were from male patients in their 70s, both residing in Nagpur. These patients approached the same state-run hospital with severe acute respiratory infection (SARI) and/or influenza-like illness (ILI) symptoms in April 2023. As per the state’s public health protocol, the hospital sent their samples to our lab for SARS-CoV-2 molecular testing, and after confirming SARS-CoV-2 with qRTPCR, these samples were used for SARS-CoV-2 genome surveillance through whole genome sequencing (WGS). Aliquots of these positive samples were subsequently used for whole genome metagenomics. However, further information about the hospitalisation, disease severity, and post-treatment progression of these patients was unavailable as they had not sought any in-patient care at that hospital.

In the metagenome composition of sample S03, the Mimiviridae family contributed 24% and 0.01% of the total virome reads and overall microbiome reads, respectively. The most abundant species within this family was *Acanthamoeba polyphaga* mimivirus, comprising 53%, followed by *Moumouvirus* at 46% and *Cafeteria roenbergensis* virus at 1% of the Mimiviridae. Other viral families were detected in minor proportions. In sample S21, the virome composition showed a similar dominance of Mimiviridae, representing 44% of the total virome and 0.1% of overall microbiome reads, respectively, with *Acanthamoeba polyphaga* mimivirus accounting for 51% within the Mimiviridae reads, followed by *Moumouvirus* at 47% and *Cafeteria roenbergensis* virus at 1%. **(Figure 1)**

**Figure 1:**
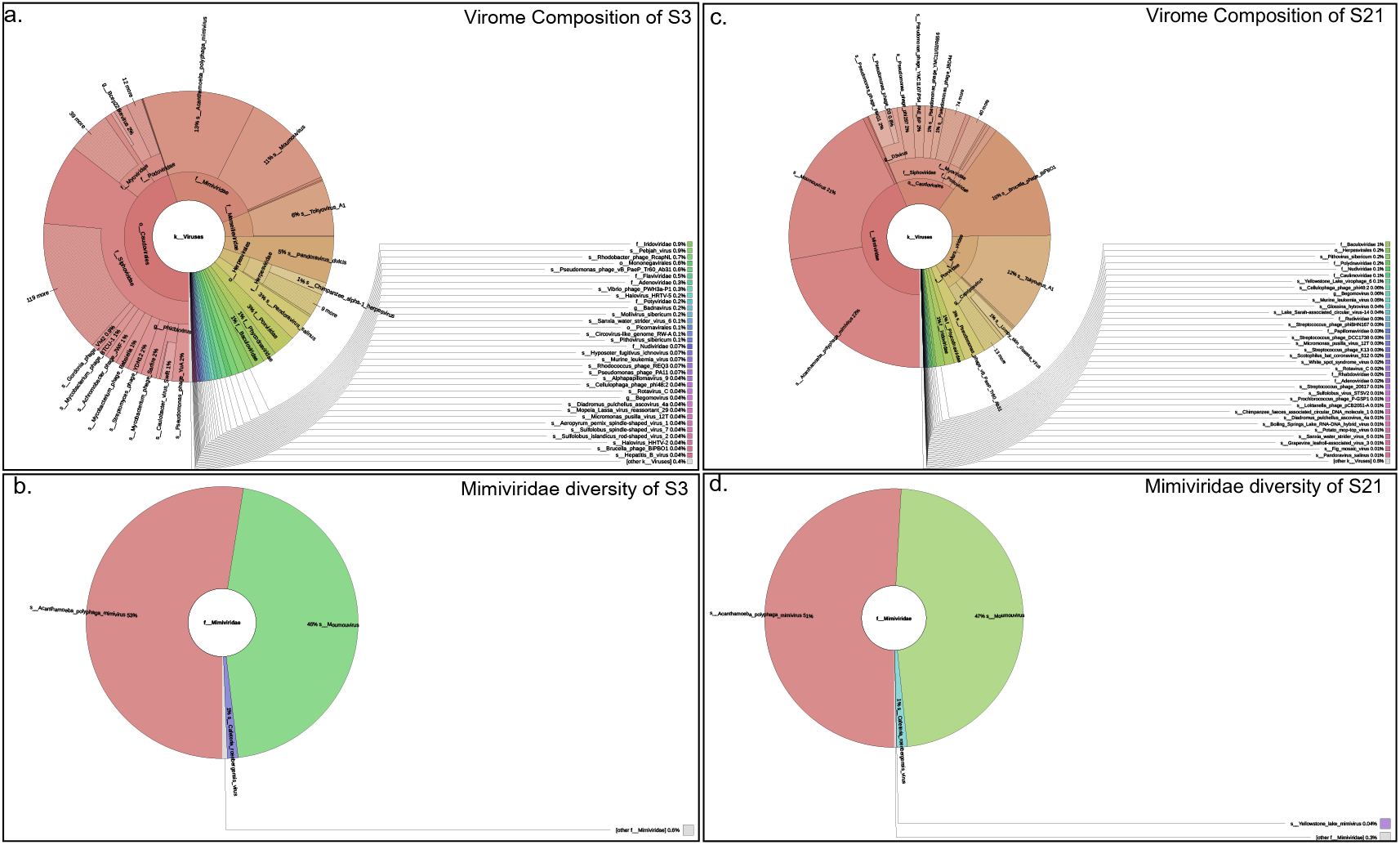
Krona Charts boxes a and c depicting the virome composition of S3 and S21, showing a predominant representation of Mimiviridae, with other notable viral families including Myoviridae, Podoviridae, and various phages. This highlights the presence of a diverse range of viral species within the sample. Boxes b and d depict the viral diversity within the Mimiviridae family.

For sample 3 **(Figure 2)**, a total of 2,031,920 reads were analyzed for generating a taxon tree, of which 15 rows of the viral taxon table passed user-defined filters (Background: None, Categories: Viruses Non-Phage, Threshold filters: Nucleotide read per million (NT rpm) >= 5, Read Specificity: All, Tree Metric: Nucleotide read counts).

**Figure 2:**
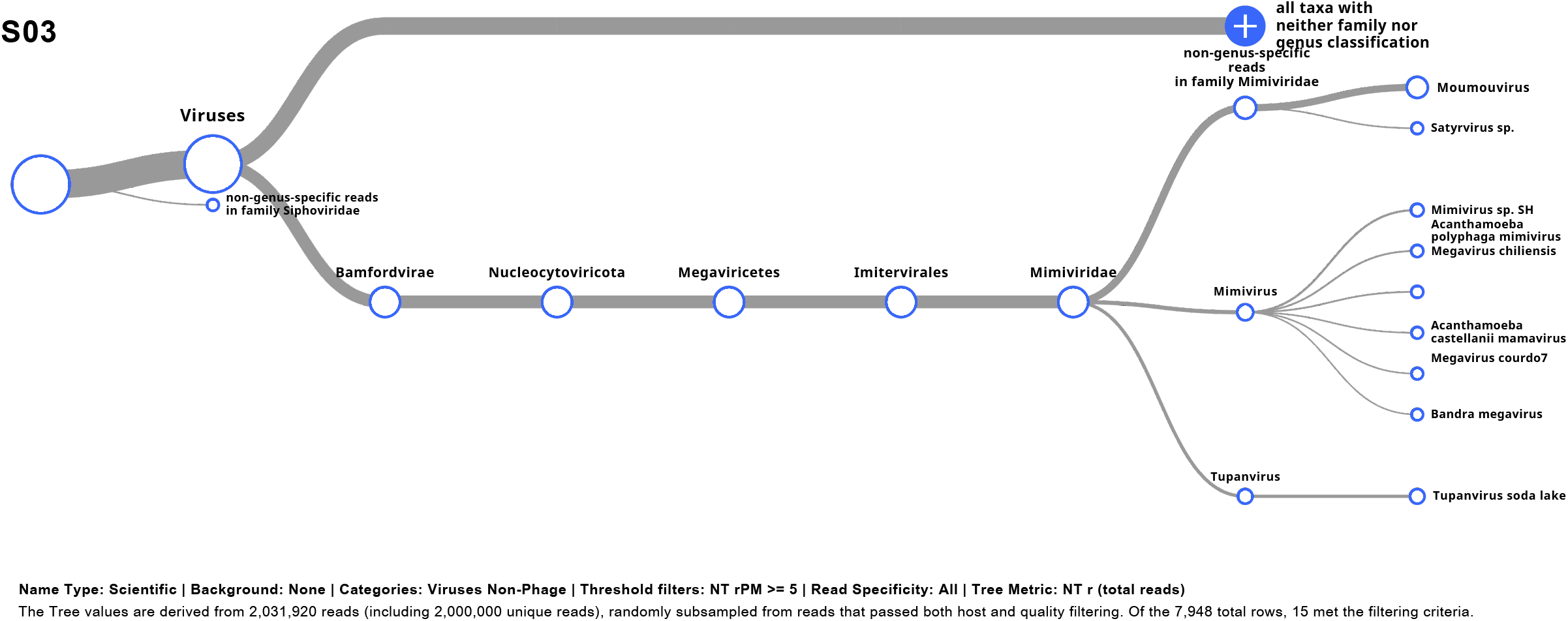
Taxonomic tree for sample S03, based on 2,031,920 reads. Mimiviridae, including *Acanthamoeba polyphaga mimivirus, Tupanvirus*, and *Moumouvirus*, along with non-genus-specific reads from Siphoviridae.

This sample shows significant members of the Mimiviridae family, including *Acanthamoeba polyphaga mimivirus, Tupanvirus soda lake*, and *Megavirus chiliensis*, along with non-genus-specific reads in the Siphoviridae family. For sample 21 **(Figure 3)**, 2,035,154 reads were processed, and 26 rows met the filtering criteria. This sample revealed additional viral taxa, including *Mimivirus U306, Megavirus vitis, Lumpy skin disease virus*, and *Cafeteria roenbergensis virus*, along with non-genus-specific reads from the Siphoviridae, Myoviridae, and Podoviridae families. The two samples collectively demonstrate the broad viral diversity captured, particularly within the Mimiviridae family and associated viral families

**Figure 3:**
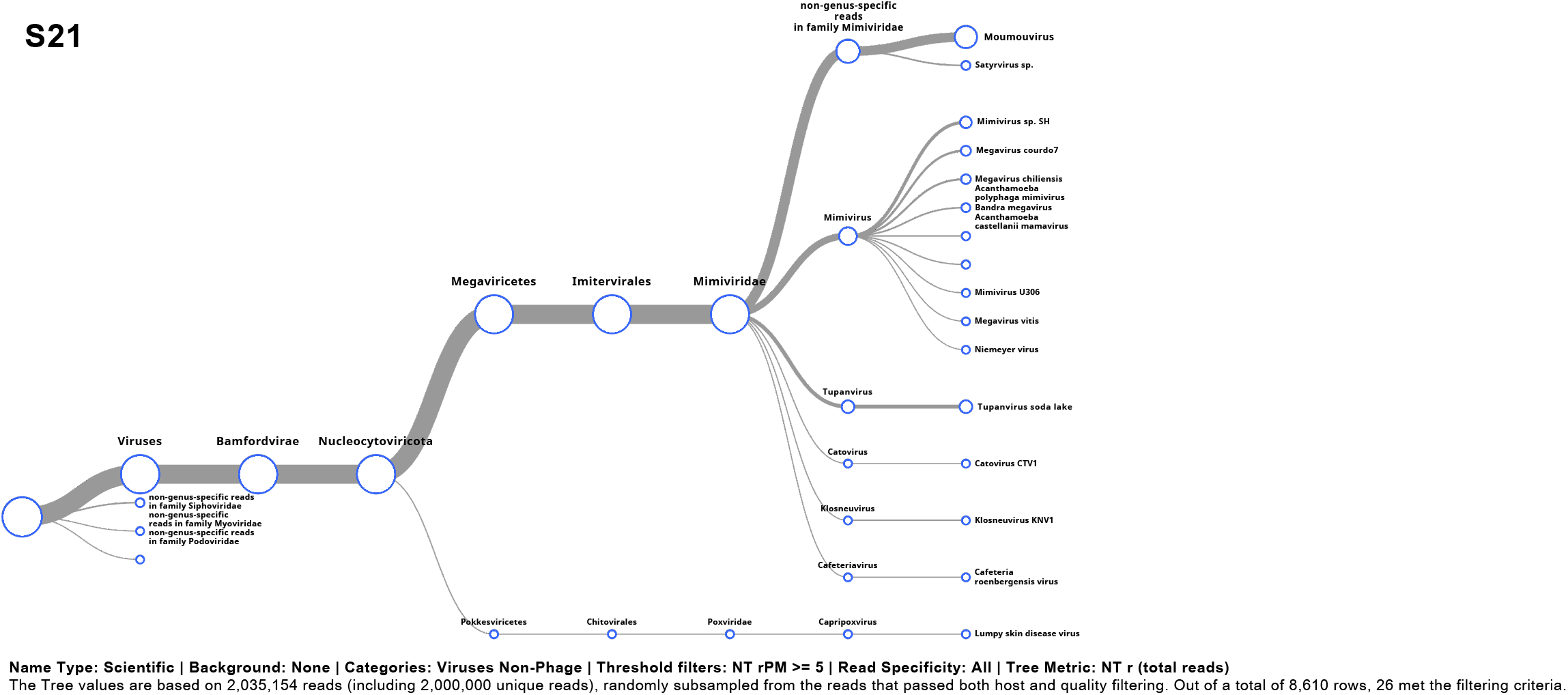
Taxonomic tree for sample S21, based on 2,035,154 reads. Key viruses include *Mimivirus U306, Megavirus vitis*, and *Lumpy skin disease virus*, with additional non-genus-specific reads from Siphoviridae, Myoviridae, and Podoviridae.

Mimivirus and Tupanvirus are the two abundant taxa in samples S03 and S21. **Figure 4** (boxes a and b) shows the coverage metrics for Mimivirus in samples S03 and S21, respectively. In both samples, consistently low viral genome coverage was observed. **Figure 4** (boxes c and d) shows the coverage metrics for Tupanvirus in the same samples. Like Mimivirus, Tupanvirus too exhibits lower genome coverage in S03 (box c) and S21 (box d).

**Figure 4:**
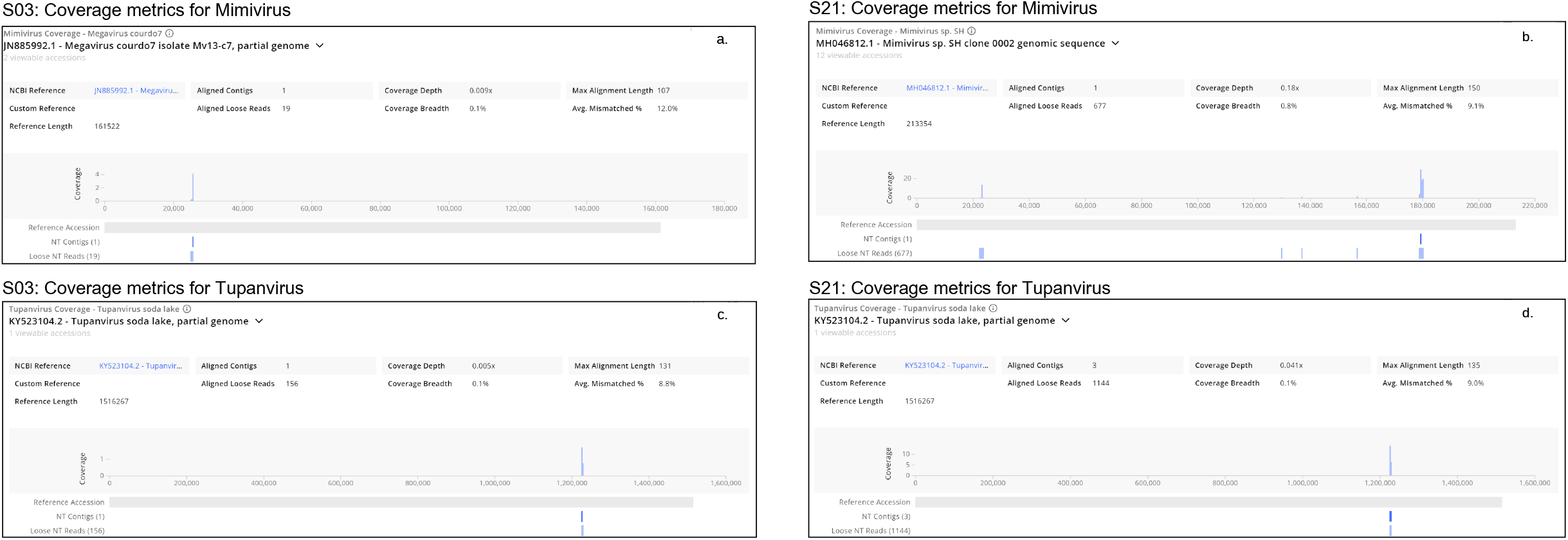
Coverage metrics for Mimivirus and Tupanvirus in samples S03 and S21.

## 4. Discussion

The detection of Mimiviridae reads in two elderly SARS-CoV-2 patients with SARI/ILI highlights a significant but underexplored aspect of viral co-occurrence in human disease. While these giant DNA viruses are primarily known to infect amoebae, their presence in the upper respiratory tract of humans, particularly in the context of co-infection with SARS-CoV-2, raises important questions about their potential role in human health and disease.

### Implications of Co-Occurrence of Giant Viruses in Respiratory Samples

The presence of such giant DNA viruses may indicate that the human respiratory system, under certain conditions, can harbor a broader diversity of viruses than commonly assumed. Although these viruses are not classically associated with human diseases, their detection in respiratory samples warrants further investigation into whether they could play a role as opportunistic pathogens, particularly in immuno-compromised individuals, such as in cases of primary infections like COVID-19.

Given that both patients were elderly, a population typically more vulnerable to severe respiratory infections, several reports suggest that Mimiviridae could exacerbate or complicate respiratory illnesses^18,19,20^. However, the exact mechanisms through which giant viruses belonging to the Mimiviridae family could influence disease severity remain unknown ^25^. However, few studies have reported Mimiviridae influencing the host immune response ^26^ or their interaction with other viral or microbial pathogen^27,28^

### Future Research Directions

The clinical relevance of Mimiviridae in respiratory infections needs to be clarified. More extensive studies will be required to assess the baseline prevalence of these viruses in different populations, particularly in patients with respiratory conditions such as ILI/SARI. Additionally, controlled studies should be conducted to determine whether these viruses are normal respiratory tract flora or contribute to disease severity through co-infections or secondary infections. Another critical area of research would be investigating the environmental reservoirs^29,30^ and transmission routes of Mimiviridae in humans. As these viruses are known to thrive in water and soil environments, understanding how they enter the human respiratory system could provide critical insights into the mechanism of their interaction with humans. One of the possible routes of Mimiviridae in humans could be through Mimiviridae hosts, such as free-living amoeba from the environment.

Given that viruses from the Mimiviridae family are commonly found in water and soil environments, their presence in the URT could be due to the patient’s consumption of contaminated water or food. The development of advanced metagenomic techniques that can more accurately detect and quantify low-abundance viral taxa like Mimivirus and Tupanvirus would be beneficial ^31,32^. Improvements in bioinformatics tools and refinement of thresholds for viral read detection could help address the challenges of low genome coverage and enhance the reliability of viral identification in clinical and environmental samples ^33,34^.

### Limitations and Challenges

Despite the intriguing nature of these findings, several limitations must be considered. The low genome coverage was observed for Mimiviridae reads in both samples. While their detection was confirmed using stringent filtering criteria, the low abundance of viral reads means we cannot definitively conclude that these viruses were actively replicating in the patients. The lack of detailed clinical follow-up data for these patients limits our ability to draw correlations between the presence of Mimiviridae and disease outcomes. Future studies should aim to integrate detailed clinical data with metagenomic findings to better understand the role of viral co-infections.

## Conclusion

Detecting Mimivirus and Tupanvirus in SARS-CoV-2-positive patients opens up new questions about the potential interaction between large DNA viruses and human health, particularly regarding respiratory infections. While the clinical significance of these viruses remains unclear, their presence in the respiratory tract, especially in vulnerable populations, underscores the need for further investigation. Future research should focus on the epidemiology, pathogenicity, and transmission dynamics of Mimiviridae in humans, as well as the development of more robust diagnostic tools to better characterize their role in co-infections and disease outcomes.

## Acknowledgement

The authors are thankful to CSIR-NEERI for providing funds under project OLP-57 (March 2023 -April 2024) for conducting this study

## Data availability statement

The details for the samples in this study are available as **“MIMIVIRUS_RAW_DATA”**

## Author declaration

The author assures that the research has followed all ethical guidelines and received approvals from the Institutional Ethics Committee for Research on Human Subjects (IEC) of CSIR-NEERI, Nagpur-20, India. Necessary consent from patients/participants has been obtained, and relevant institutional documentation has been archived.

## Author contribution statement

SST and KK have contributed equally to the conceptualization, experimentation, and data analysis of this study.

